# Injustices in pandemic vulnerability: A spatial-statistical analysis of the CDC Social Vulnerability Index and COVID-19 outcomes in the U.S.

**DOI:** 10.1101/2021.05.27.21257889

**Authors:** Jamie Song, Eugenia South, Sara Solomon, Douglas Wiebe

## Abstract

**Background:** The COVID-19 pandemic has exacerbated health injustices in the U.S. driven by racism and other forms of structural violence. Research has shown the disproportionate impacts of COVID-19 morbidity and mortality in the most marginalized communities.

**Objectives:** We examined the associations between COVID-19 cumulative incidence (CI) and case-fatality risk (CFR) and the CDC’s Social Vulnerability Index (SVI), a composite score assessing historical marginalization and thus vulnerability to disaster events.

**Methods:** Using county-level data from national databases, we used population density, Gini index, percent uninsured, and average annual temperature as covariates, and employed negative binomial regression to evaluate relationships between SVI and COVID-19 outcomes. Optimized hot spot analysis identified hot spots of COVID-19 CI and CFR, which were compared in terms of SVI using logistic regression.

**Results:** As of 2/3/21, 26,452,031 cases of and 448,786 deaths from COVID-19 had been reported in the U.S. Negative binomial regression showed that counties in the top SVI quintile reported 13.7% higher CI (p<0.001) than those in the bottom SVI quintile. Additionally, each unit increase in a county’s SVI score was associated with a 0.2% increase in CFR (p<0.001). Logistic regression analysis showed that counties in the lowest SVI quintile had significantly greater odds of being in a CI hot spot than all other counties, yet counties in the highest SVI quintile had 63% greater odds (p=0.008) of being in a CFR hot spot than counties in the lowest SVI quintile.

**Conclusion:** We demonstrated a significant relationship between SVI and CFR, but the relationship between SVI and CI is complex and warrants further investigation. SVI may help elucidate unequal impacts of COVID-19 and guide prioritization of vaccines to communities most impacted by structural injustices.

## Background

The COVID-19 pandemic has exacerbated existing health inequities around the world, and especially in highly impacted nations like the U.S. As of February 3rd, 2021, 26,452,031 confirmed cases of COVID-19 and 448,786 deaths attributable to COVID-19 had been reported in all 50 U.S. states, putting the U.S. at the top in case count and death toll among all countries.^1^ These cases and deaths have disproportionately burdened the most marginalized communities – those that have borne the brunt of racism and white supremacy, capitalism and economic domination, and other dimensions of structural violence embedded in U.S. society.^2^ This marginalization has led to drastic differences in the abilities of communities to prevent, mitigate, and recover from outbreaks of COVID-19.

For instance, mortality rates among people who are Black, Indigenous, Latine,^3^ and Pacific Islander have exceeded that of white people; in particular, Black people have died at 1.4 times the rate of white people.^4^ The disproportionate impact of COVID-19 is also laid bare in the Centers for Disease Control and Prevention (CDC)’s latest life expectancy estimates from the first half of 2020: while life expectancy declined by 1 year overall, it dropped by 2.7 years among Black people and by 1.9 years among Latine people.^a^ The growing life expectancy gap between white Americans and people of color, particularly those who are Black and Latine, is already alarming, and expected to worsen after analysis of data from a tumultuous second half of 2020. This inequity reflects underlying differences in vulnerability that are constituted by a deeply rooted history of U.S. white supremacy, which has manifested in structural racism, classism, and other forms of domination that generate health injustices.

The CDC Social Vulnerability Index (SVI) is a composite metric that assesses forms of historical marginalization and thus vulnerability to the impacts of disaster events. It is composed of 15 individual variables organized into four “themes,” which are Socioeconomic Status (Theme 1), Household Composition & Disability (Theme 2), Minority Status & Language (Theme 3), and Housing Type & Transportation (Theme 4).^6^ While SVI has received growing attention as a potential tool for guiding health equity efforts amidst the COVID-19 pandemic, few studies have validated the use of SVI for this purpose. To address this gap, this study examined the associations between COVID-19 outcome variables, namely cumulative incidence (CI) and case fatality risk (CFR),^b^ and the SVI, along with its component themes. Validating the potential use of SVI to understand critical inequities relevant to COVID-19 may provide support for its use as a public health tool, including the use of SVI to guide prioritization of COVID-19 vaccination efforts.

## Methods

An ecological study assessing the relationship between the SVI and COVID-19 outcome variables (CI and CFR) was conducted at the county level within the contiguous United States (48 states). Data sources included the CDC, Johns Hopkins University, NOAA, and the U.S. Census Bureau, and analyses involved spatial statistics as well as regression models. Geospatial “optimized hot spot analysis” (OHSA) was used to identify broader areas of impact for COVID-19 CI and CFR; in order to preserve the fidelity of the OHSA, Alaska and Hawaii were excluded from this study. Stata IC 16.1 was used for statistical analysis. ArcMap 10.7 was used for geospatial analysis.

### Study Sample

County-level Social Vulnerability Index (SVI) scores were obtained from the Centers for Disease Control and Prevention (CDC)’s Agency for Toxic Substances and Disease Registry (ATSDR).^8^ Publicly available county-level COVID-19 data (on CI and CFR) were downloaded from the Johns Hopkins University (JHU) Center for Systems Science and Engineering (CSSE) Github repository.

Potential covariates were also ascertained for this study. Demographic data for all U.S. counties were downloaded from the American Community Survey (ACS) database.^9^ Data on average temperature over 12 months (February 2020 to January 2021) by U.S. county were obtained from the U.S. National Oceanic and Atmospheric Administration (NOAA) National Centers for Environmental Information (NCEI). A shapefile of 2018 U.S. county boundaries was obtained from the U.S. Census Bureau.^10^ Ultimately, 3,081 counties (and other local geographies that are operationally defined as counties, e.g. “cities” in the Commonwealth of Virginia) were included in this study. Most counties in the State of Utah,^c^ as well as the counties of Dukes and Nantucket in the Commonwealth of Massachusetts, were excluded at the geospatial mapping stage due to missing COVID-19 outcome data. At the point of statistical analysis, the County of Rio Arriba in the State of New Mexico was excluded due to missing SVI data.

### Measures

The SVI was developed by Flanagan and colleagues in response to the field of disaster management’s primary focus on physical hazards and the historical lack of consideration of social factors that affect impact.^11^ It is a composite metric of “social vulnerability,” which is defined by Flanagan and colleagues in terms of a community’s attributes that affect its capacity to “anticipate, confront, repair, and recover” from disaster events.^11^ The SVI has been applied to research on COVID-19 in the U.S.,^12,13,14^ and has been adapted into other metrics such as Snyder and Parks’ socio-ecological vulnerability index,^15^ and the Pandemic Vulnerability Index developed for the NIH by Marvel and colleagues.^16^ In this study, we used the SVI and its component “themes” as proxies for the various forms of social, economic, and political marginalization that are historically embedded and spatially heterogeneous across the U.S. The CDC ATSDR is now responsible for maintaining the SVI database and conducting biannual updates of the dataset. In this study, we utilize the latest update of the dataset, which was released in March 2020 and reflects data estimates from 2018.^8^ The SVI and its component themes are percentile scores with values ranging between 0 and 100.

The Johns Hopkins University (JHU) Center for Systems Science and Engineering (CSSE) has maintained one of the most robust databases of COVID data in the country, which was used for this study. The database provides daily counts of COVID-19 confirmed cases and deaths for geographic areas around the world, as well as CI per 100,000 persons (coded as Incidence Rate) and CFR (coded as Case Fatality Ratio). For the U.S., the JHU CSSE database includes data at the county level as well as data unassigned to county-level geographic areas, but only the former were used in this study.

The American Community Survey (ACS) is conducted by the U.S. Census Bureau on an ongoing basis to collect vital demographic data about the United States.^9^

Covariates were selected for possible association with COVID-19 outcomes and evaluated for multicollinearity prior to incorporation into regression analyses. These included data from the ACS on 1) population density (operationally defined as persons per square mile), the Gini coefficient of income inequality (a single number measuring how much a county’s income distribution deviates from an equal distribution), and 3) percentage of population without health insurance, as well as data on 4) average county temperature (over 12 months) from NOAA.

### Geospatial and Statistical Analysis

Choropleth maps were used to visualize COVID-19 CI and CFR across counties (Fig. 1), as well as SVI (Fig. 2), by quintile. Quadratic prediction plots were used to visualize the linearity of relationships between COVID-19 outcome variables (CI and CFR) and overall SVI score, as well as the SVI’s four ‘themes’ (Fig. 3).

Potential covariates were evaluated for multicollinearity using the variance inflation factor (VIF) test prior to inclusion in analysis. The VIF measures the correlation and strength of correlation between the explanatory variables in a regression model; VIF values over 5 generally indicate potentially severe correlation between explanatory variables, which would entail exclusion of the covariate in question.

To check for linearity, likelihood-ratio tests were conducted for each relationship to compare models using raw SVI (linear model) and those using SVI quintiles (nonlinear model). For those relationships determined to be linear, negative binomial regression was used to model the relationships between COVID-19 outcome variables and overall SVI scores, as well as SVI theme scores (Table 1a). For relationships determined to be significantly nonlinear, negative binomial regression was performed between the outcome variables and overall SVI quintiles, as well as SVI theme quintiles (Table 1b). All analyses were conducted with the four ascertained covariates.

Getis-Ord Gi* hot spot analysis is a geostatistical approach used to identify statistically significant spatial clusters of high values (hot spots) and low values (cold spots). Optimized hot spot analysis (OHSA) is a tool that further interrogates the dataset and determines the specific model parameters that will produce optimal results. In this study, OHSA was used to identify geospatially significant hot and cold spots of COVID-19 CI and CFR at the 95% confidence level (Fig. 4). The demographic characteristics of counties in hot and cold spots were summarized (Table 2). Independence between counties in CI and CFR hot and cold spot groups was tested using Pearson’s χ^2^ test of independence (Table 3). Associations between hot spot status and SVI were then evaluated using logistic regression (Table 4).

## Results

### Descriptive statistics and linearity tests

As of 02/03/21, 26,089,070 cases of and 443,214 deaths from COVID-19 had been reported in the 3,081 U.S. counties represented in this study. 362,961 more cases and 5,572 more deaths had been reported in the U.S. at this point, but were excluded either because they were in Alaska or Hawaii or because they were unassigned to specific county-level geographic areas.

When all covariates underwent the VIF test with overall SVI as well as its component themes, VIF values for each covariate were below 2, indicating no significant multicollinearity. Likelihood-ratio tests demonstrated that the relationship between CI and SVI was nonlinear (LR χ^2^=37.50, df=3, p<0.0001), as well as the relationship between CI and SVI Theme 1 (LRχ^2^=30.56, df=3, p<0.0001). These analyses also showed that the relationship between CI hot spot status and overall SVI was nonlinear (LR χ^2^=28.41, df=3, p<0.0001), as well as the relationships between CI hot spot status and Theme 1 (LR χ^2^=60.84, df=3, p<0.0001) and CI hot spot status and Theme 3 (LR χ^2^=17.67, df=3, p=0.0005). Relationships between CFR hot spot status and SVI Themes 2 (LR χ^2^=11.20, df=3, p=0.0107), 3 (LR χ^2^=9.40, df=3, p=0.0244), and 4 (LR χ^2^=11.49, df=3, p=0.0094) were also nonlinear, and the relationship between CFR hot spot status and overall SVI was nearly nonlinear (LR χ^2^=7.15, df=3, p=0.0674). All other relationships were determined to be linear (p>0.05).

Negative binomial regression models with covariates showed that counties in the top SVI quintile experienced 21% higher CI (p<0.001) than counties in the bottom quintile (Table 1a). Counties in the middle (3rd) quintile for SVI Theme 1 (Socioeconomic Status) experienced 5.1% lower CI (p=0.013) than those in the bottom quintile, while counties in the top quintile experienced 6.0% greater CI (p=0.019) than those in the bottom quintile. Among linear relationships, each unit increase in SVI Theme 2 (Housing Composition & Disability) was associated with a 0.063% increase in CI (p=0.009); each unit increase in SVI Theme 3 (Minority Status & Language) was associated with a 0.065% increase in CI (p=0.014); and each unit increase in SVI Theme 4 (Housing Type & Transportation) was associated with a 0.086% increase in CI (p=0.001) (Table 1b).

Each unit increase in a county’s overall SVI score as well as scores in Themes 1-2 was associated with an increase in CFR of 0.20-0.35% (p<0.001) (See Table 1b). However, a one unit increase in SVI Theme 3 was associated with a 0.23% decrease in CFR (p<0.001). There was no significant county-level relationship between SVI Theme 4 and CFR (p=0.261).

### Geospatial analysis

OHSA generated CI hot spots (N=864), CI cold spots (N=784), CFR hot spots (N=902), and CFR cold spots (N=1,212). Compared to the overall population, Black, Indigenous, and white people were overrepresented in CI hot spots, whereas Asian/Pacific Islander, white, and foreign-born people were overrepresented in CI cold spots; in CFR hot spots, Black, Latine, and foreign-born people were overrepresented, while Asian/Pacific Islander and white people were overrepresented in CFR cold spots (Table 2). Pearson’s χ^2^ test of independence demonstrated that CI and CFR hot and cold spots were significantly independent (p=0.007) (Table 3).

Logistic regression models controlling for covariates demonstrated a largely negative relationship between SVI and CI hot spot status (Table 4a). Counties in the lowest SVI quintile had significantly greater odds (ranging from 56.4% to 120% [p<0.006]) of being in a CI hot spot compared to all other counties. The relationship between SVI Theme 1 (Socioeconomic Status) and CI hot spot status was similar; counties in the lowest quintile for Theme 1 also had significantly greater odds (ranging from 88.5% to 213% [p<0.001]) of being in a CI hot spot compared to all other counties. Counties in the middle quintile had 35.9% greater odds (p=0.018) of being in a CI hot spot compared to counties in the lowest quintile. Additionally, each one unit increase in a county’s SVI Theme 4 (Housing Type and Transportation) score was associated with a 0.81% decrease in its odds of being in a CI hot spot (p<0.001) (Table 4b).

Counties in the highest SVI quintile had 63% greater odds (p=0.008) of being in a CFR hot spot than counties in the lowest quintile (Table 4a). Additionally, counties in the top three quintiles of SVI Theme 3 (Minority Status and Language) had at least 138% greater odds (p<0.003) of being in a CFR hot spot than counties in the lowest quintile, and counties in the top quintile of SVI Theme 4 (Housing and Transportation) had 47% greater odds (p=0.013) of being in a CFR hot spot than counties in the lowest quintile. In contrast, counties in the lowest quintile of SVI Theme 2 (Household Composition and Disability) had significantly greater odds (ranging from 38.8% to 66.4% [p<0.025]) of being in a CFR hot spot than all other counties. No significant relationship was found between SVI Theme 1 (Socioeconomic Status) and CFR hot spot status (p=0.183).

## Discussion

In this study, we examined the associations between COVID-19 outcomes, namely cumulative incidence (CI) and case-fatality risk (CFR), and the CDC SVI, along with its component themes. We used both spatial and non-spatial models with covariates to describe a complex relationship between SVI and CI, but a clearer, positive relationship between SVI and CFR. The elucidation of nonlinear relationships between SVI and COVID-19 outcomes challenges existing literature on SVI and COVID-19 that has assumed or failed to question linearity in this relationship.^17,18^ Other results, including the strong correspondence between Theme 3 (Minority Status & Language) and CFR hot spot status, confirm and extend existing conclusions in previous work.^14,17,19^ Methodologically, we brought optimized hot spot analysis (OHSA) to bear on county-level case and death counts, and attempted to clarify broader regions of high COVID-19 impact as well as correct for reporting bias in counties with underreported cases and/or deaths. This spatial-statistical analysis of COVID-19 outcomes invites further investigations to employ spatially aware methods that attempt to account for the spatiotemporal nature of infectious disease epidemiology.

We found that counties in the top quintile for overall SVI experienced significantly greater CI than those in the bottom quintile, and similarly, those in the top quintile for Theme 1 (Socioeconomic Status) experienced greater CI than those in the bottom SVI quintile. We also discovered positive linear associations between SVI Themes 2, 3, and 4 and CI. These relationships may be partially explained by the increased crowded living conditions of people living in counties with higher SVI. They may also be employed in riskier labor sectors, such as manufacturing, service, and health care. Older age distributions as well as greater prevalence of chronic health conditions in more marginalized communities may facilitate progression of SARS-CoV-2 infection to the clinical stage, increasing the likelihood of seeking diagnostic testing. However, a largely negative association between SVI and CI hot spot status was also observed. We propose that lower vulnerability is also associated with factors that facilitate SARS-CoV-2 transmission, including the ability to travel more frequently.^20^ We urge further research studying the relationship between SVI and CI, perhaps using a time series analysis or possibly more granular data that are not available to the public, either of which could help elucidate more micro-level phenomena.

Increases in overall SVI, as well as in SVI Themes 1 and 2, were strongly associated with increased COVID-19 CFR. This positive relationship was validated by the association between SVI and CFR hot spot status. Residents of counties with high SVI may experience greater prevalence of chronic health conditions that have been identified as comorbidities increasing the risk of mortality following SARS-CoV-2 infection,^21^ including cancer, respiratory illness, cardiovascular illness, and diabetes.^22,23,24^ These populations may also have poorer access to health care, especially the sophisticated medical care that is often required to mitigate severe and mortal COVID-19 outcomes. Furthermore, we observed an association of even greater magnitude between SVI Theme 3 (Minority Status & Language) and CFR, suggesting that communities of color and/or immigrant communities affected by COVID-19 may face greater risks of severe illness and mortality than white communities. This disparity reflects known health injustices imposed on communities of color through structural racism.^25^

In a descriptive analysis of demographics in CI and CFR hot and cold spots we found that, compared to the general population of the U.S., Indigenous people were overrepresented in CI hot spots, Latine people were overrepresented in CFR hot spots, and Black people were overrepresented in both.

We also found that COVID-19 CI hot and cold spots were significantly different from CFR hot and cold spots, meaning that the clustered areas with the greatest and least CI had little spatial overlap with those with the greatest and least CFR. Therefore, preventive measures, mitigation strategies, and recovery efforts must be prioritized differently based on an area’s incidence and mortality risks.

Ultimately, our results suggest that our most marginalized communities, including BIPOC (Black, Indigenous, and people of color) and working class people, are also the most vulnerable to COVID-19 mortality. As health equity work in federal, state, and local governments as well as in communities across the U.S. endeavors to focus resources and support on people and neighborhoods most vulnerable to COVID-19 impact, tools like the SVI as well as geospatial and statistical analysis can help identify and target those vulnerabilities.

### Limitations

We identified several limitations of our study. First, we relied on national data reported by thousands of localities, which inevitably introduced bias. Many cases and deaths were geographically non-specific, or specific to geographies that were not classified under the U.S. county system. For example, the majority of the state of Utah was not represented in this study because the reported data were not specific to county-level geographies. Furthermore, our results are likely biased by underreporting of cases by localities and states; in addition, we were unable to account for disparities in testing that would differentially deflate incidence estimates. We likely also underestimated mortality, based on studies showing significant underreporting of COVID-19 mortality in the U.S.^26^

Secondly, our county-level analysis is subject to ecological bias; observations of county-level CI and CFR are unable to fully capture individual or even community-level disease transmission phenomena. This bias could have been reduced with data at the ZIP code or census tract levels, which were not publicly available because of ethical and welfare concerns with COVID-19 data privacy.

Third, the variable sizes of U.S. counties may have negatively impacted the validity of Getis-Ord Gi* hot spot analysis,^27^ though the optimization algorithms performed in OHSA may have helped to account for this problem.

Fourth, SVI is a composite metric, and it does have advantages in assessing many different dimensions of marginalization or vulnerability, but because we only used the overall index and component subthemes, we were unable to clarify the effects of individual variables.

Fifth, our study controlled for population density, income inequality, foreign-born population, and lack of health insurance as covariates, but we did not assess interaction effects that may represent sources of confounding. Further research may explore the differential moderating effects of these variables on the relationship between SVI and CI as well as CFR.

Sixth, our study analyzed COVID-19 data from January 23^rd^, 2020 to February 3^rd^, 2021.The lack of a temporal analysis over such a long period limits this study’s ability to assess the variable influence of SVI on COVID-19 outcomes over time. CFR may be more significantly biased by the lack of temporal analysis, since best practices to manage COVID and reduce poor outcomes in clinical settings were developed later in the pandemic. The confounding effect of time in this case could be mitigated by a time series analysis, or even a mixed effects model incorporating time as a random effect. Future investigators may also consider a space-time cube analysis, a tool that can elucidate areas that experienced persistent COVID-19 impact, which may be more strongly associated with SVI.

## Conclusion

In this ecological study, we found that SVI shows significant positive associations with COVID-19 CI at a county level, yet it is negatively associated with being in a CI hot spot. We also showed that SVI is significantly associated with COVID-19 county-level case fatality as well as CFR hot spot status. Overall, we suggest that social vulnerability is more strongly associated with COVID-19 case severity and fatality than it is with incidence, implying that more marginalized communities tend to experience greater traumas and human losses.

This study validates the use of SVI in current vaccine prioritization policy. We recommend the continued application of SVI in vaccine distribution in tandem with other factors, such as race and age, to address the structural racism, class domination, and other injustices that enforce gaps in health equity. We also urge key decision makers in public policy to use SVI and/or other metrics of marginalization in prioritizing programs for COVID-19 recovery and pandemic resilience, which should entail behavioral health care access, neighborhood economic development, and direct reparations to those harmed by historical traumas and injustices, including people who are Black and/or Indigenous.

## Data Availability

I am currently developing a plan for making my original dataset available. In the meantime, I can be reached at for any data requests.

## Acknowledgments

The authors would like to thank Dr. Robert Schnoll for his guidance on manuscript preparation, study design, and methodology, as well as his support throughout the Capstone II seminar; Dr. E. Paul Wileyto for his guidance on statistical methods; Vicky Tam for her guidance on geospatial methods and study limitations; Dr. Hillary Nelson and Heather Klusaritz for their support and mentorship throughout the Capstone I seminar; and Jonathan Delgadillo for his assistance on regression model methodology in Stata.

## Footnotes

a Note: We use “Latine” in this study instead of similar terms like “Latino/a” and “Latinx”. See reference 5: Katie Slemp, “Latino, Latina, Latin@, Latine, and Latinx: Gender Inclusive Oral Expression in Spanish” (2020). *Electronic Thesis and Dissertation Repository*. 7297. https://ir.lib.uwo.ca/etd/7297

b Note: For CFR, we use “risk” instead of “rate” or “ratio.” See reference 7: Kelly H, Cowling BJ. Case Fatality: Rate, Ratio, or Risk? *Epidemiology*. 2013;24(4):622-623. doi:10.1097/EDE.0b013e318296c2b6

c These counties were: Beaver, Box Elder, Cache, Carbon, Daggett, Garfield, Grand, Iron, Juab, Kane, Millard, Morgan, Piute, Rich, Sevier, Uintah, Washington, Wayne, and Weber.

## Notes

### Competing Interest Statement

The authors have declared no competing interest.

### Funding Statement

No funding was received for this project.

### Author Declarations

No IRB necessary

