## Appendix and Supplemental Material for "Injustices in pandemic vulnerability: A spatial-statistical analysis of the CDC Social Vulnerability Index and COVID-19 outcomes in the U.S."

Figure 1. Quintile and hot spot map of COVID-19 cumulative incidence (CI; above) and COVID-19 case fatality risk (CFR; below) by U.S. county, Feb. 3, 2020

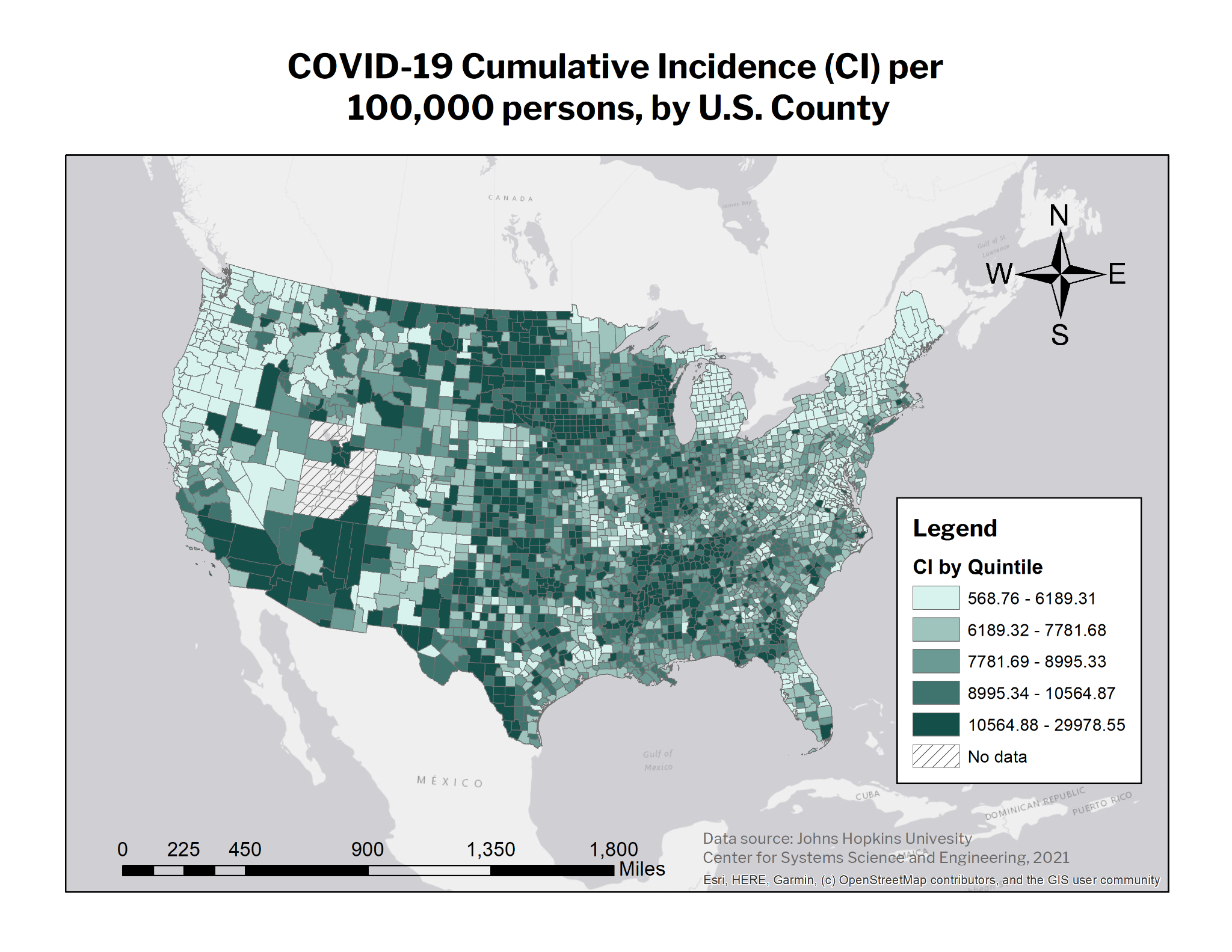

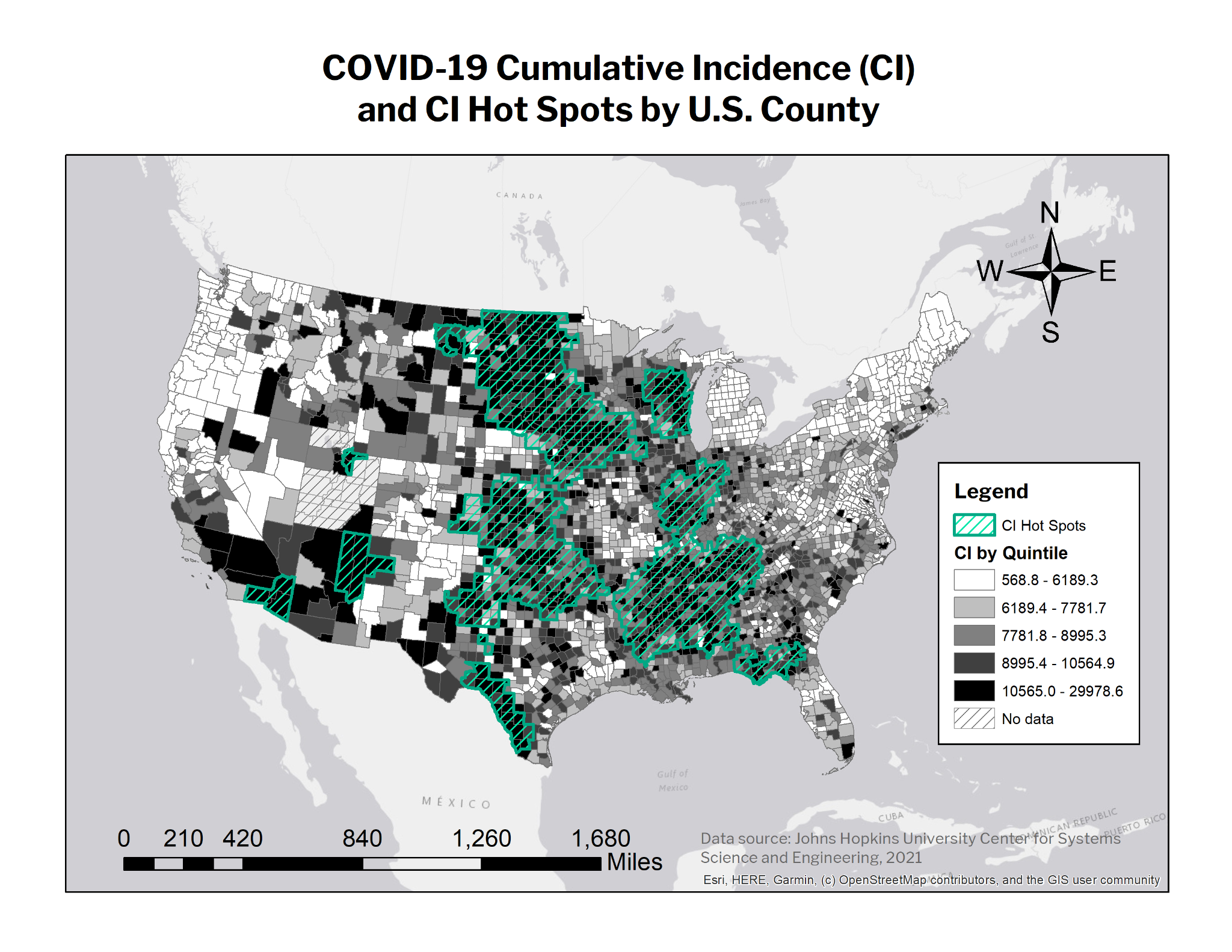

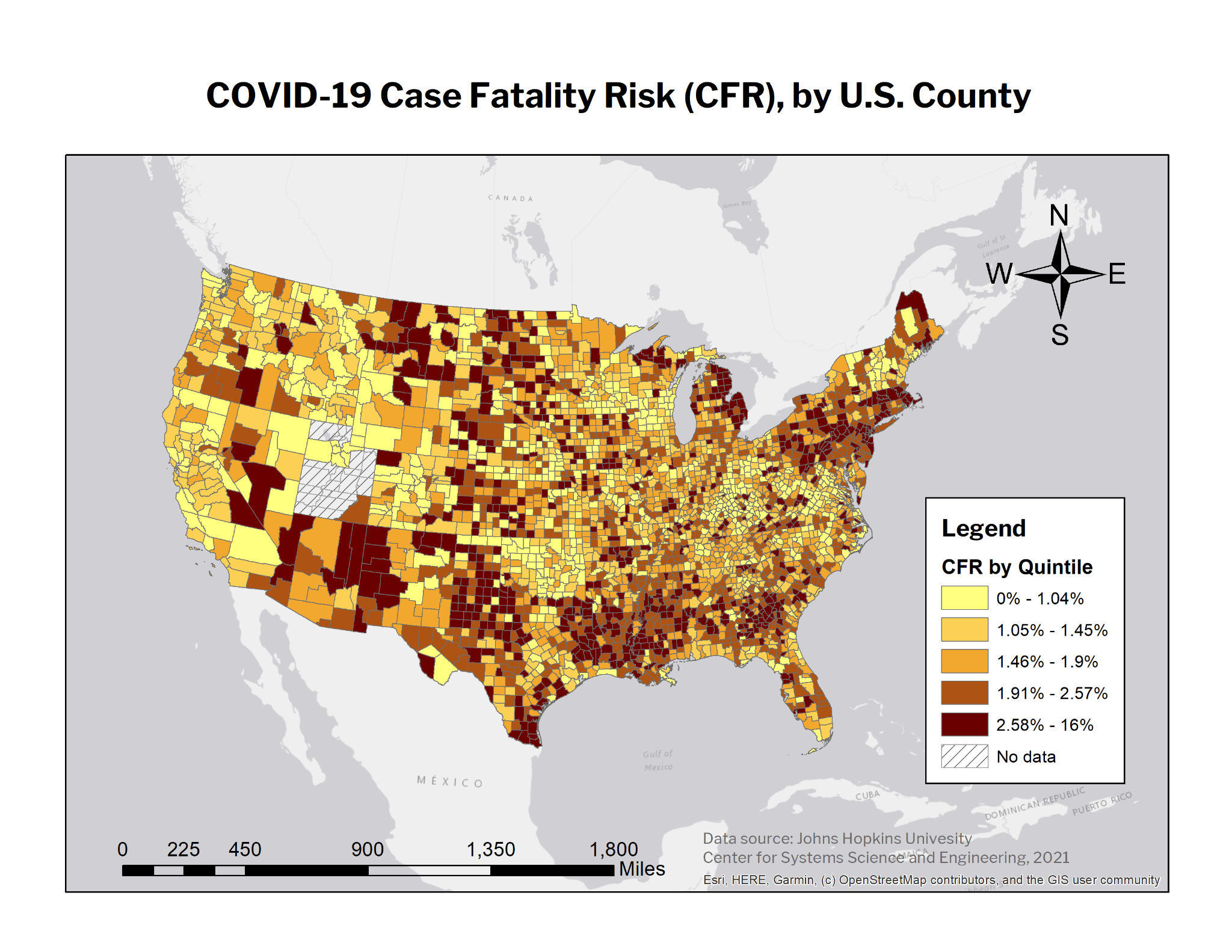

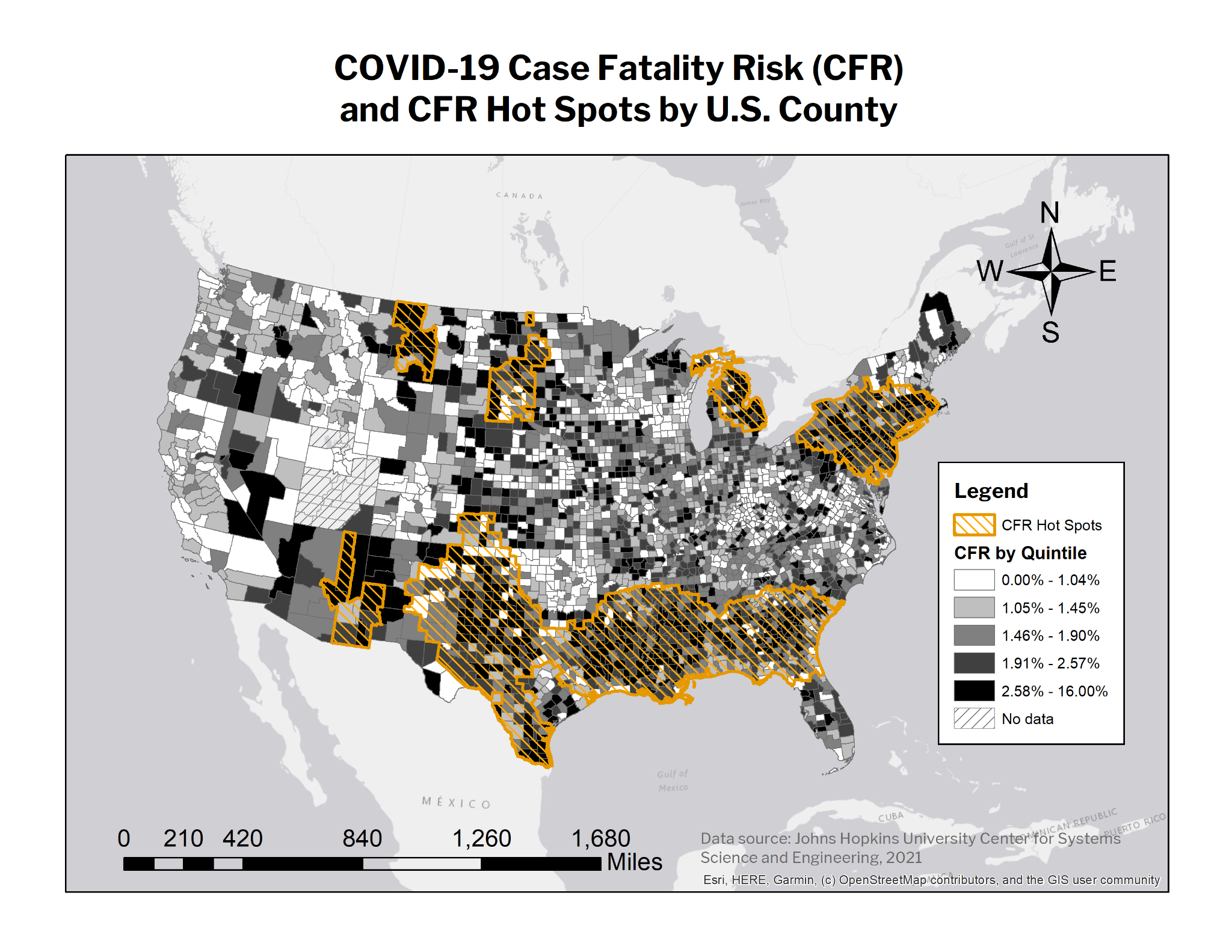

Figure 2. Quintile map of SVI scores by U.S. county (2018), with COVID-19 CI and CFR hot spots superimposed

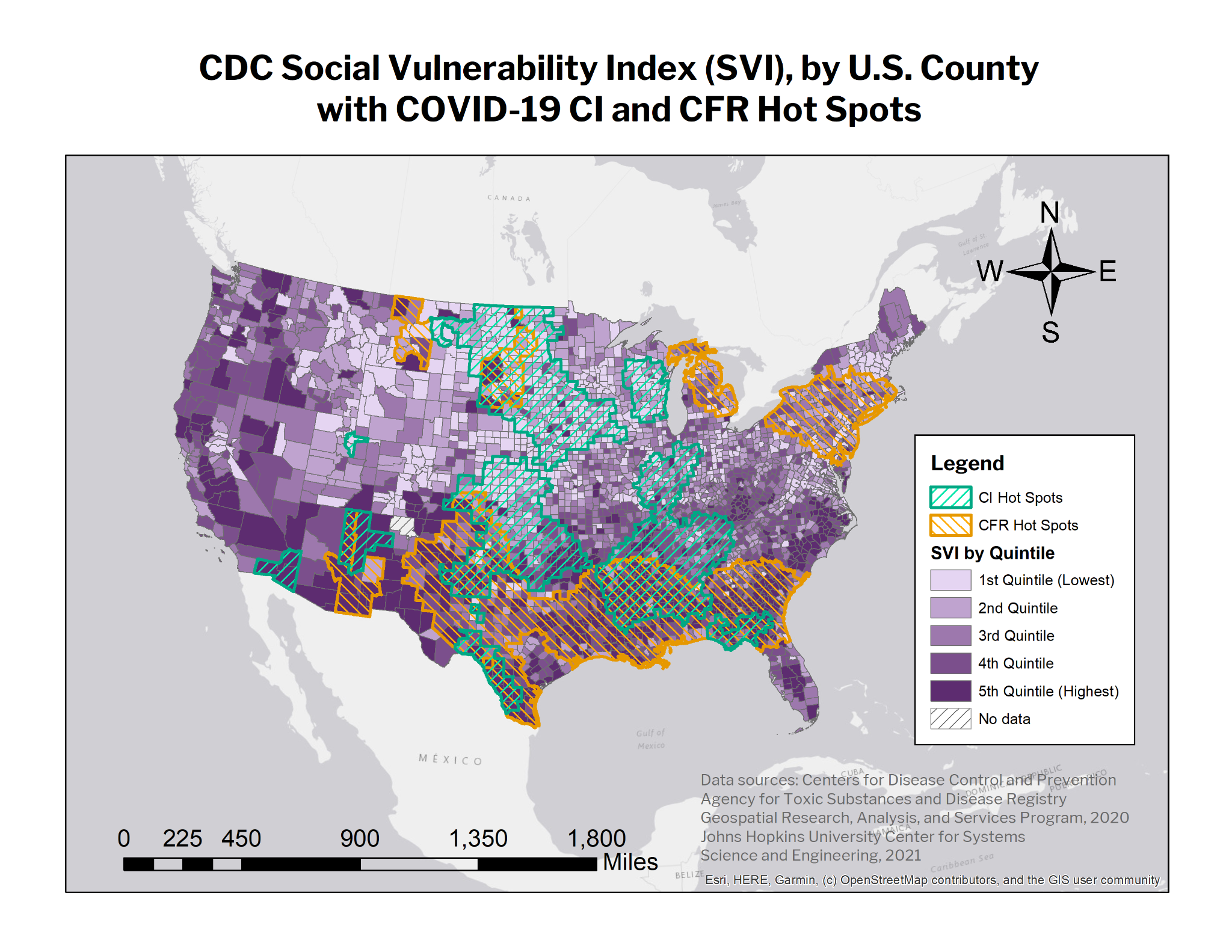

Table 1. Results of negative binomial regression analyses between overall SVI & SVI themes and COVID-19 outcome variables (with covariates)

1a. Results for nonlinear relationships

| ***Overall SVI vs. Cases*** | **Incidence rate ratio (IRR)** | **p-value** | **95% confidence interval** |
| --- | --- | --- | --- |
| *1st quintile* |  |  |  |
| *2nd* | 0.9909093 | 0.653 | (0.952278, 1.031108) |
| *3rd* | 0.9703354 | 0.160 | (0.9304415, 1.01194) |
| *4th* | 1.01995 | 0.390 | (0.9750589, 1.066908) |
| *5th* | 1.137486 | <0.001 | (1.080707, 1.197248) |

| ***SVI Theme 1 vs. Cases*** | **Incidence rate ratio (IRR)** | **p-value** | **95% confidence interval** |
| --- | --- | --- | --- |
| *1st quintile* |  |  |  |
| *2nd* | 0.9691491 | 0.125 | (0.9311479, 1.008701) |
| *3rd* | 0.9486836 | 0.013 | (0.9098794, 0.9891427) |
| *4th* | 0.9674149 | 0.139 | (0.9258609, 1.010834) |
| *5th* | 1.060233 | 0.019 | (1.00962, 1.113382) |

1b. Results for linear relationships

| ***Cumulative Incidence (CI)*** | **Incidence Rate Ratio (IRR)** | **p-value** | **95% Confidence Interval** |
| --- | --- | --- | --- |
| ***vs. Theme 2*** | 1.000633 | 0.009 | (1.000161, 1.001105) |
| ***vs. Theme 3*** | 1.00065 | 0.014 | (1.000134, 1.001167) |
| ***vs. Theme 4*** | 1.000859 | 0.001 | (1.000365, 1.001354) |

| ***Case-Fatality Risk (CFR)*** | **Incidence Rate Ratio (IRR)** | **p-value** | **95% Confidence Interval** |
| --- | --- | --- | --- |
| ***vs. SVI*** | 1.002043 | <0.001 | (1.001186, 1.0029) |
| ***vs. Theme 1*** | 1.002912 | <0.001 | (1.002113, 1.003712) |
| ***vs. Theme 2*** | 1.00346 | <0.001 | (1.002797, 1.004125) |
| ***vs. Theme 3*** | 0.9977207 | <0.001 | (0.9969899, 0.998452) |
| ***vs. Theme 4*** | 0.9995896 | 0.261 | (0.9988747, 1.000305) |

Table 2. Population density, race, and nativity demographic characteristics of cumulative incidence (CI) and case-fatality risk (CFR) hot and cold spots in contiguous 48 U.S. states (*Numbers greater than overall are in* **bold**)

| **Demographic variables** | **CI** | | **CFR** | | ***Overall population*** *(not total)* |
| --- | --- | --- | --- | --- | --- |
|  | ***Cold Spot*** | ***Hot Spot*** | ***Cold Spot*** | ***Hot Spot*** |  |
| *Total population* | 124,307,032 | 36,474,182 | 117,439,516 | 102,412,224 | 321,614,559 |
| *Land area (sq. miles)* | 637,088.65 | 676,693.80 | 1,010,007.54 | 735,633.44 | 2,888,071.82 |
| *Population density (persons/sq. mi.)* | **195.12** | 53.90 | **116.28** | **139.22** | 111.36 |
| *Black population (% of total)* | 14,601,208 (11.7%) | 5,070,141 (**13.9%**) | 10,496,304 (8.9%) | 17,727,865 (**17.3%**) | 39,922,789 (12.4%) |
| *Indigenous population* | 425,156 (0.34%) | 620,468 (**1.7%**) | 736,307 (0.63%) | 351,799 (0.34%) | 2,035,668 (0.63%) |
| *Asian/Pacific Islander population* | 8,709,599 (**7.0%**) | 767,157 (2.1%) | 7,724,063 (**6.6%**) | 5,170,321 (5.0%) | 17,526,041 (5.4%) |
| *Latine population* | 17,444,843 (14.0%) | 3,348,174 (9.2%) | 19,079,779 (16.2%) | 18,616,034 (**18.2%**) | 58,145,508 (18.0%) |
| *White population* | 79,473,239 (**63.9%**) | 25,741,732 (**70.6%**) | 75,904,150 (**64.6%**) | 58,267,049 (56.9%) | 195,603,617 (60.8%) |
| *Foreign-born population* | 18,114,789 (**14.6%**) | 2,055,479 (5.6%) | 15,031,560 (12.8%) | 14,517,900 (**14.2%**) | 43,642,933 (13.6%) |

Table 3. Results of Pearson’s χ^2^ test of independence between COVID outcome variable hot and cold spots

|  |  | ***Case-Fatality Risk (CFR)*** | |  |
| --- | --- | --- | --- | --- |
|  |  | **Cold Spot** | **Hot Spot** | *Totals* |
| ***Cumulative Incidence (CI)*** | **Cold Spot** | 365 | 222 | *588* |
|  | **Hot Spot** | 293 | 247 | *540* |
|  | *Totals* | *658* | *469* | *1,127* |

Pearson’s χ^2^ statistic = 7.2636

p = 0.007

Table 4. Results of logistic regression analyses between overall SVI & SVI themes and COVID-19 outcome variable hot spot status (with covariates)

4a. Results for nonlinear relationships

| **Overall SVI vs. CI hot spot status** | **Odds ratio (OR)** | **p-value** | **95% confidence interval** |
| --- | --- | --- | --- |
| *1st quintile*  *(comparison group)* |  |  |  |
| *2nd* | 0.6366347 | <0.001 | (0.4950553, 0.818704) |
| *3rd* | 0.4653587 | <0.001 | (0.3540531, 0.611656) |
| *4th* | 0.4546024 | <0.001 | (0.3400616, 0.6077232) |
| *5th* | 0.6393726 | 0.006 | (0.4642218, 0.8806078) |

| **SVI Theme 1 vs. CI hot spot status** | **Odds ratio (OR)** | **p-value** | **95% confidence interval** |
| --- | --- | --- | --- |
| *1st quintile*  *(comparison group)* |  |  |  |
| *2nd* | 0.5305544 | <0.001 | (0.4108377, 0.6851563) |
| *3rd* | 0.319499 | <0.001 | (0.241138, 0.4233246) |
| *4th* | 0.3567157 | <0.001 | (0.2676767, 0.4753724) |
| *5th* | 0.5056991 | <0.001 | (0.3706724, 0.6899128) |

| **SVI Theme 3 vs. CI hot spot status** | **Odds ratio (OR)** | **p-value** | **95% confidence interval** |
| --- | --- | --- | --- |
| *1st quintile = comparison group* |  |  |  |
| *2nd* | 1.305616 | 0.033 | (1.021642, 1.668523) |
| *3rd* | 1.358742 | 0.018 | (1.054163, 1.751323) |
| *4th* | 1.105561 | 0.476 | (0.8391795, 1.4565) |
| *5th* | 0.7848907 | 0.132 | (0.5728672, 1.075386) |

| **Overall SVI vs. CFR hot spot status** | **Odds ratio (OR)** | **p-value** | **95% confidence interval** |
| --- | --- | --- | --- |
| *1st quintile = comparison group* |  |  |  |
| *2nd* | 1.007505 | 0.965 | (0.7237439, 1.402521) |
| *3rd* | 1.117471 | 0.507 | (0.8050427, 1.551149) |
| *4th* | 1.100106 | 0.578 | (0.7862891, 1.53917) |
| *5th* | 1.632877 | 0.008 | (1.139513, 2.339847) |

| **SVI Theme 2 vs. CFR hot spot status** | **Odds ratio (OR)** | **p-value** | **95% confidence interval** |
| --- | --- | --- | --- |
| *1st quintile = comparison group* |  |  |  |
| *2nd* | 0.6008435 | 0.001 | (0.4468585, 0.807891) |
| *3rd* | 0.6783993 | 0.008 | (0.5086152, 0.90486) |
| *4th* | 0.6467233 | 0.003 | (0.4858629, 0.8608415) |
| *5th* | 0.7206135 | 0.024 | (0.5420568, 0.9579877) |

| **SVI Theme 3 vs. CFR hot spot status** | **Odds ratio (OR)** | **p-value** | **95% confidence interval** |
| --- | --- | --- | --- |
| *1st quintile = comparison group* |  |  |  |
| *2nd* | 1.676853 | 0.003 | (1.189126 2.364625) |
| *3rd* | 2.419313 | <0.001 | (1.741938 3.360093) |
| *4th* | 2.379985 | <0.001 | (1.708586 3.315215) |
| *5th* | 2.414106 | <0.001 | (1.710312 3.407512) |

| **SVI Theme 4 vs. CFR hot spot status** | **Odds ratio (OR)** | **p-value** | **95% confidence interval** |
| --- | --- | --- | --- |
| *1st quintile = comparison group* |  |  |  |
| *2nd* | 1.158977 | 0.328 | (1.189126 2.364625) |
| *3rd* | 0.8555172 | 0.317 | (1.741938 3.360093) |
| *4th* | 1.045676 | 0.773 | (1.708586 3.315215) |
| *5th* | 1.472546 | 0.013 | (1.710312 3.407512) |

4b. Results for linear relationships

| ***CI hot spot status*** | **Odds Ratio (OR)** | **p-value** | **95% Confidence Interval** |
| --- | --- | --- | --- |
| ***vs. Theme 2*** | 1.00285 | 0.076 | (0.9996975, 1.006012) |
| ***vs. Theme 4*** | .991938 | <0.001 | (0.9888292, .9950565) |

| ***CFR hot spot status*** | **Odds Ratio (OR)** | **p-value** | **95% Confidence Interval** |
| --- | --- | --- | --- |
| ***vs. Theme 1*** | 1.002599 | 0.183 | (0.9987786, 1.006434) |

**Supplemental Materials**

Table A. Results of likelihood-ratio tests for linearity

A1. Likelihood-ratio tests for **negative binomial regressions** to test linearity of relationships between SVI and COVID-19 outcome variables (including covariates)

| ***Cumulative Incidence (CI)*** | **Likelihood-ratio chi2 (3)** | **p-value** | **Linearity** |
| --- | --- | --- | --- |
| ***vs. SVI*** | 37.50 | <0.0001 | Nonlinear |
| ***vs. Theme 1*** | 30.56 | <0.0001 | Nonlinear |
| ***vs. Theme 2*** | 2.19 | 0.5338 | Linear |
| ***vs. Theme 3*** | 2.58 | 0.4610 | Linear |
| ***vs. Theme 4*** | 0.40 | 0.9406 | Linear |

| ***Case-Fatality Risk (CFR)*** | **Likelihood-ratio chi2 (3)** | **p-value** | **Linearity** |
| --- | --- | --- | --- |
| ***vs. SVI*** | 2.24 | 0.5238 | Linear |
| ***vs. Theme 1*** | 4.05 | 0.2561 | Linear |
| ***vs. Theme 2*** | 3.25 | 0.3543 | Linear |
| ***vs. Theme 3*** | 1.71 | 0.6342 | Linear |
| ***vs. Theme 4*** | 5.02 | 0.1702 | Linear |

A2. Likelihood-ratio tests for **logistic regressions** to test linearity of relationships between SVI and COVID-19 outcome variable hot spot status (including covariates)

| ***CI Hot Spot Status*** | **Likelihood-ratio chi2 (3)** | **p-value** | **Linearity** |
| --- | --- | --- | --- |
| ***vs. SVI*** | 28.41 | <0.0001 | Nonlinear |
| ***vs. Theme 1*** | 60.84 | <0.0001 | Nonlinear |
| ***vs. Theme 2*** | 1.97 | 0.5776 | Linear |
| ***vs. Theme 3*** | 17.67 | 0.0005 | Nonlinear |
| ***vs. Theme 4*** | 3.95 | 0.2674 | Linear |

| ***CFR Hot Spot Status*** | **Likelihood-ratio chi2 (3)** | **p-value** | **Linearity** |
| --- | --- | --- | --- |
| ***vs. SVI*** | 7.15 | 0.0674 | Nearly nonlinear |
| ***vs. Theme 1*** | 0.25 | 0.9699 | Linear |
| ***vs. Theme 2*** | 11.20 | 0.0107 | Nonlinear |
| ***vs. Theme 3*** | 9.40 | 0.0244 | Nonlinear |
| ***vs. Theme 4*** | 11.49 | 0.0094 | Nonlinear |

Table B. Regression model attributes

B1. Negative binomial regression models

|  |  | **Pseudo R^2^** | **LR χ^2^ (df=1)*** | **p-value** |
| --- | --- | --- | --- | --- |
| ***Cumulative Incidence (CI)*** | ***vs. SVI*** | 0.0032 | 1.9e6 | <0.001 |
|  | ***vs. Theme 1*** | 0.0028 | 1.9e6 | <0.001 |
|  | ***vs. Theme 2*** | 0.0022 | 2.0e6 | <0.001 |
|  | ***vs. Theme 3*** | 0.0022 | 2.1e6 | <0.001 |
|  | ***vs. Theme 4*** | 0.0023 | 2.0e6 | <0.001 |
| ***Case Fatality Risk (CFR)*** | ***vs. SVI*** | 0.0060 | 6.7e4 | <0.001 |
|  | ***vs. Theme 1*** | 0.0071 | 6.6e4 | <0.001 |
|  | ***vs. Theme 2*** | 0.0090 | 6.5e4 | <0.001 |
|  | ***vs. Theme 3*** | 0.0066 | 6.9e4 | <0.001 |
|  | ***vs. Theme 4*** | 0.0053 | 6.8e4 | <0.001 |

*Note: This likelihood-ratio test compares the negative binomial model and the Poisson model. A p-value of less than 0.05 indicates that the negative binomial model is significantly more appropriate due to overdispersion in the outcome variable.

B2. Logistic regression models

|  |  | **Pseudo R^2^** |
| --- | --- | --- |
| ***Cumulative Incidence (CI)*** | ***vs. SVI*** | 0.0447 |
|  | ***vs. Theme 1*** | 0.0550 |
|  | ***vs. Theme 2*** | 0.0339 |
|  | ***vs. Theme 3*** | 0.0384 |
|  | ***vs. Theme 4*** | 0.0401 |
| ***Case Fatality Risk (CFR)*** | ***vs. SVI*** | 0.1729 |
|  | ***vs. Theme 1*** | 0.1698 |
|  | ***vs. Theme 2*** | 0.1730 |
|  | ***vs. Theme 3*** | 0.1795 |
|  | ***vs. Theme 4*** | 0.1737 |
